# Genetic variants are identified to increase risk of COVID-19 related mortality from UK Biobank data

**DOI:** 10.1101/2020.11.05.20226761

**Authors:** Jianchang Hu, Cai Li, Shiying Wang, Ting Li, Heping Zhang

## Abstract

**Background:** The severity of coronavirus disease 2019 (COVID-19) caused by the severe acute respiratory syndrome coronavirus 2 (SARS-CoV-2) is highly heterogenous. Studies have reported that males and some ethnic groups are at increased risk of death from COVID-19, which implies that individual risk of death might be influenced by host genetic factors.

**Methods:** In this project, we consider the mortality as the trait of interest and perform a genome-wide association study (GWAS) of data for 1,778 infected cases (445 deaths, 25.03%) distributed by the UK Biobank. Traditional GWAS failed to identify any genome-wide significant genetic variants from this dataset. To enhance the power of GWAS and account for possible multi-loci interactions, we adopt the concept of super-variant for the detection of genetic factors. A discovery-validation procedure is used for verifying the potential associations.

**Results:** We find 8 super-variants that are consistently identified across multiple replications as susceptibility loci for COVID-19 mortality. The identified risk factors on Chromosomes 2, 6, 7, 8, 10, 16, and 17 contain genetic variants and genes related to cilia dysfunctions (*DNAH7* and *CLUAP1*), cardiovascular diseases (*DES* and *SPEG*), thromboembolic disease (*STXBP5*), mitochondrial dysfunctions (*TOMM7*), and innate immune system (*WSB1*). It is noteworthy that *DNAH7* has been reported recently as the most downregulated gene after infecting human bronchial epithelial cells with SARS-CoV2.

**Conclusions:** Eight genetic variants are identified to significantly increase risk of COVID-19 mortality among the patients with white British ancestry. These findings may provide timely evidence and clues for better understanding the molecular pathogenesis of COVID-19 and genetic basis of heterogeneous susceptibility, with potential impact on new therapeutic options.

## Introduction

Coronavirus disease 2019 (COVID-19) is a highly infectious disease caused by the severe acute respiratory syndrome coronavirus 2 (SARS-CoV-2). The pneumonia was first reported in December 2019 in Wuhan, Hubei Province, China, followed by an outbreak across the country [1, 2]. As of September 8th, 2020, the pandemic of COVID-19 has rapidly spread worldwide and caused over 27 million infected cases and 891,000 deaths (3.3%) according to JHU COVID-19 dashboard [3]. Currently, the effective therapeutic measures available to counteract the SARS-CoV-2 are limited. While studies have been dedicated to investigating the clinical features, epidemiological characteristics of COVID-19 [4-11], and genomic characterization of SARS-CoV-2 [12], few are through the lens of statistical genetics and the host genetic factors contributing to COVID-19 remain largely enigmatic [13, 14]. Moreover, the severity of COVID-19 and course of the infection is highly heterogenous. The majority of COVID-19 cases only have mild or no symptoms, while some of the patients develop serious health outcomes. A UK cross-sectional survey of 20,133 patients who were hospitalized with COVID-19 showed that patients with diabetes, cardiovascular diseases, hypertension, or chronic respiratory diseases were at higher risk of death [15]. More importantly, evidence has shown that males and some ethnic groups have increased risk of death from COVID-19 [16-20]. These observations suggest that there might be host genetic determinants which predispose the subgroup of patients to more severe COVID-19 outcomes. Undoubtedly, there is an urgent need for understanding host genetic basis of heterogeneous susceptibility to COVID-19 and uncovering genetic risk factors. Current studies mainly focus on investigating associations between host genetic factors and infection or respiratory failure [13, 14]. Obviously, infection may only be partially explained by genetic factors since exposure to the virus could be more important. Here, we consider the mortality as the trait of interest for our analysis.

As of early August 2020, UK Biobank [21, 22] has released the testing results of COVID-19 for 12,428 participants, including 1,778 (14.31%) infected cases with 445 deaths related to COVID-19. This dataset accompanied by already available health care data, genetic data and death data offers a unique resource and timely opportunity for learning the host genetic determinants of COVID-19 susceptibility, severity, and mortality.

In this project, we perform a genome-wide association study (GWAS) exploiting the concept of super-variates in statistical genetics to identify potential risk loci contributing to the COVID-19 mortality. A super-variant is a combination of alleles in multiple loci in analogue to a gene. However, in contrast to a gene that refers to a physically connected region of a chromosome, the loci contributing to a super-variant is not restricted by its spatial location in the genome [23-25]. The rationale behind our analysis is two-fold: First, COVID-19 infections require environmental exposure and the genetic contribution may be limited relative to the environmental exposure, and the mortality may have a stronger genetic effect. Second, COVID-19 is a complex syndrome, which may reflect interacting genomic factors, and our analysis with super-variants enables leveraging gene interactions beyond the additive effects.

## Methods

### Sample processing and genotype quality control

We analyze the COVID-19 data released by UK Biobank (Category ID: 100091) [22] on August 3rd, 2020, which include in total 1,778 of COVID-19 infected cases. Here, we consider an infected case as a sample with any positive PCR test result or a death with virus found. Among infected cases, 445 of them were reported death caused directly or indirectly by COVID-19 and the remainder of 1333 patients are survivors. In our analysis, to limit the potential effect of population structure, we focus on samples from white British ancestry. After standard sample quality controls, there remain 1096 of COVID-19 infected participants, of which 292 were deaths (26.64%) and 804 were survivors. Their imputed genotype data (Field ID: 22801-22822) and clinical variables including gender and age (Field ID: 31, 34) are all accessible from UK Biobank [21].

Our analysis makes use of imputed single-nucleotide polymorphism (SNP) datasets from UK Biobank. SNPs with duplicated names and positions are excluded. After standard genotyping quality control, where variants with low call rate (missing probability ≥ 0.05) and disrupted Hardy-Weinberg equilibrium (p-value < 1×10^−6^) are removed, we retain in total 18,617,478 SNPs. We divide the whole SNP dataset into 2734 non-overlapping local sets according to the physical position so that each set consists of SNPs within a segment of physical length 1 Mb.

### Statistical analysis

We consider the concept of super-variant for GWAS. A super-variant is a combination of alleles in multiple loci, but unlike a gene that refers to a physically connected region of chromosome, the loci contributing to a super-variant can be anywhere in the genome [24, 25]. The super-variant is suggested to be powerful and stable in association studies as it aggregates the strength of individual signals. In addition, it accounts for potential complex interactions between different genes even when they are located remotely. To identify significant super-variants, a local ranking and aggregation method is adopted. Chromosomes are divided into local SNP sets. Within each set, random forest technique is utilized to obtain the so-called depth importance measure of each SNP which leads to a ranking of SNPs in terms of their importance. Top SNPs within each local set are then aggregated into a super-variant. In addition, two modes of transmission, dominant and recessive modes are both considered for the super-variant identification. We refer the readers to [25] for details.

Our analysis considers the following discovery-validation procedure. The complete dataset is randomly divided into two sets, one for discovery and the other for verification. Each set consists of 146 deaths and 402 survivors. We apply the aforementioned ranking and aggregation method for super-variant identification on the discovery dataset. After the discovery of the super-variants, we then investigate their associations with the death outcomes of COVID-19 through logistic regression in the verification and complete datasets. Age and gender are considered in the regression analyses as confounders to remove potential bias. We use 1.83×10^−5^ (i.e., 0.05/2734) as the threshold for super-variant-level association on the discovery dataset since 2,734 SNP sets are considered. A super-variant is verified if its logistic regression coefficient achieves the level of 0.05 significance on the verification dataset and super-variant-level significance on the complete dataset.

To ascertain the stability of the associations, we repeat the above procedure for 10 times, and retain the verified super-variants and their contributing SNPs. Finally, for super-variants that are consistently verified across multiple runs, we conduct Cox regressions with adjustment for age and gender in the complete dataset to further validate their associations.

## Results

We find 216 different verified super-variants across 10 repetitions of the discovery-validation procedure. More importantly, there are two super-variants, chr6_148 and chr7_23, identified in 4 out of 10 repetitions. In addition, there are 6 super-variants, chr2_197, chr2_221, chr8_99, chr10_57, chr16_4 and chr17_26 identified in 3 out of 10 repetitions. According to the binomial distribution, the probability of a super-variant being verified in 4 (3) out of 10 repetitions by chance is at most 0.00096 (0.0105) if p-value in the verification dataset is assumed to be uniformly distributed.

In terms of the SNPs contributing to these 8 super-variants, there exist SNPs selected multiple times across different repetitions. Specifically, for chr6_148, SNP rs117928001 is a contributing SNP in all 4 times when this super-variant is verified, and there are other 94 contributing SNPs selected 3 times. Similarly, for chr7_23, SNP rs1322746 is a contributing SNP in 3 repetitions when this super-variant is verified, and other 4 SNPs are selected 2 times. For super-variant chr2_197 which is identified in 3 out of 10 repetitions, SNPs rs34011564 and rs71040457 are both contributing SNPs in all 3 times. For chr8_99, SNPs rs4735444 and rs531453964 are contributing SNPs of verified super-variants in all 3 repetitions. SNPs rs117217714, rs2176724, rs9804218 and rs2301762 are contributing SNPs for chr17_26, chr2_197, chr10_57 and chr16_4 in all 3 repetitions when these super-variants are verified, respectively. We calculate minor allele frequency (MAF), odds ratio (OR), and p-value for the contributing SNPs of the 8 super-variants based on the complete dataset. See Table S1 in Additional file 1 for the details of all contributing SNPs which are selected in at least 2 repetitions.

We use SNPs which are selected in at least 2 repetitions to representatively form 8 super-variants according to the same mode of transmission (dominant/recessive) when they are discovered. Table 1 gives their effects estimated from univariate logistic regression and Cox regression with adjustment for sex and age in the complete dataset. For the logistic regression, all of them achieve super-variant-level significance (i.e., p-value < 1.83×10^−5^). The strongest signal in terms of p-value is given by chr7_23 (p-value = 9.5×10^−9^), and the largest odds ratio appears at chr17_26 (OR = 4.237). For the Cox regression, the largest individual hazards ratio (HR) appears at chr17_26 (HR = 2.956) as well, and the smallest individual p-value is given by chr2_221 (p-value = 5.2×10^−9^). Table 2 lists the details of representative contributing SNPs with high selection frequency and important gene mapping results of the 8 super-variants. Figure 1 shows that the survival probabilities of the patients with identified super-variants remarkably drop during the first 20 days since testing, suggesting of risk genotypes. Figure 2 presents the survival probabilities stratified by the number of super-variants. The HR of super-variants is 1.778 with 95% CI being [1.593, 1.985], and the associated p-value is 1.1×10^−24^, while the p-values of sex and age are 1.2×10^−2^ (HR = 1.489, male) and 2.9×10^−18^ (HR = 1.107), respectively. The survival probability of patients with more than 3 super-variants dramatically decreases to around 0.6 during the first three weeks.

**Table 1:** Marginal effects of 8 super-variants in the complete dataset.

| **Table 1 \| Marginal effects of 8 super-variants in the complete dataset.** |  |  |  |  |  |  |  |
| --- | --- | --- | --- | --- | --- | --- | --- |
| Dominant | Gene | OR | 95% CI of OR | p value | HR | 95% CI of HR | p value |
| chr6_148 | *STXBP5/STXBP5-AS1* | 2.909 | [1.938, 4.365] | 1.4x10<sup>-7</sup> | 2.048 | [1.435, 2.921] | 7.7x10<sup>-5</sup> |
| chr8_99 | *CPQ* | 1.923 | [1.419, 2.605] | 1.6x10<sup>-5</sup> | 1.502 | [1.119, 2.015] | 6.7x10<sup>-3</sup> |
| chr16_4 | *CLUAP1* | 2.725 | [1.744, 4.259] | 7.0x10<sup>-6</sup> | 2.123 | [1.433, 3.143] | 1.7x10<sup>-4</sup> |
| chr17_26 | *WSB1* | 4.237 | [2.472, 7.263] | 8.4x10<sup>-8</sup> | 2.956 | [1.949, 4.482] | 3.4x10<sup>-7</sup> |
| Recessive | Gene | OR | 95% CI of OR | p value | HR | 95% CI of HR | p value |
| ch2_197 | *DNAH7/SLC39A10* | 2.553 | [1.801, 3.616] | 7.3x10<sup>-8</sup> | 1.625 | [1.170, 2.257] | 3.8x10<sup>-3</sup> |
| chr2_221 | *DES/SPEG* | 2.739 | [1.893, 3.963] | 4.9x10<sup>-8</sup> | 2.614 | [1.894, 3.609] | 5.2x10<sup>-9</sup> |
| chr7_23 | *TOMM7* | 2.411 | [1.774, 3.276] | 9.5x10<sup>-9</sup> | 1.943 | [1.451, 2.603] | 8.1x10<sup>-6</sup> |
| chr10_57 | *PCDH15* | 2.521 | [1.736, 3.662] | 7.1x10<sup>-7</sup> | 1.813 | [1.283, 2.561] | 7.4x10<sup>-4</sup> |

**Table 2:**
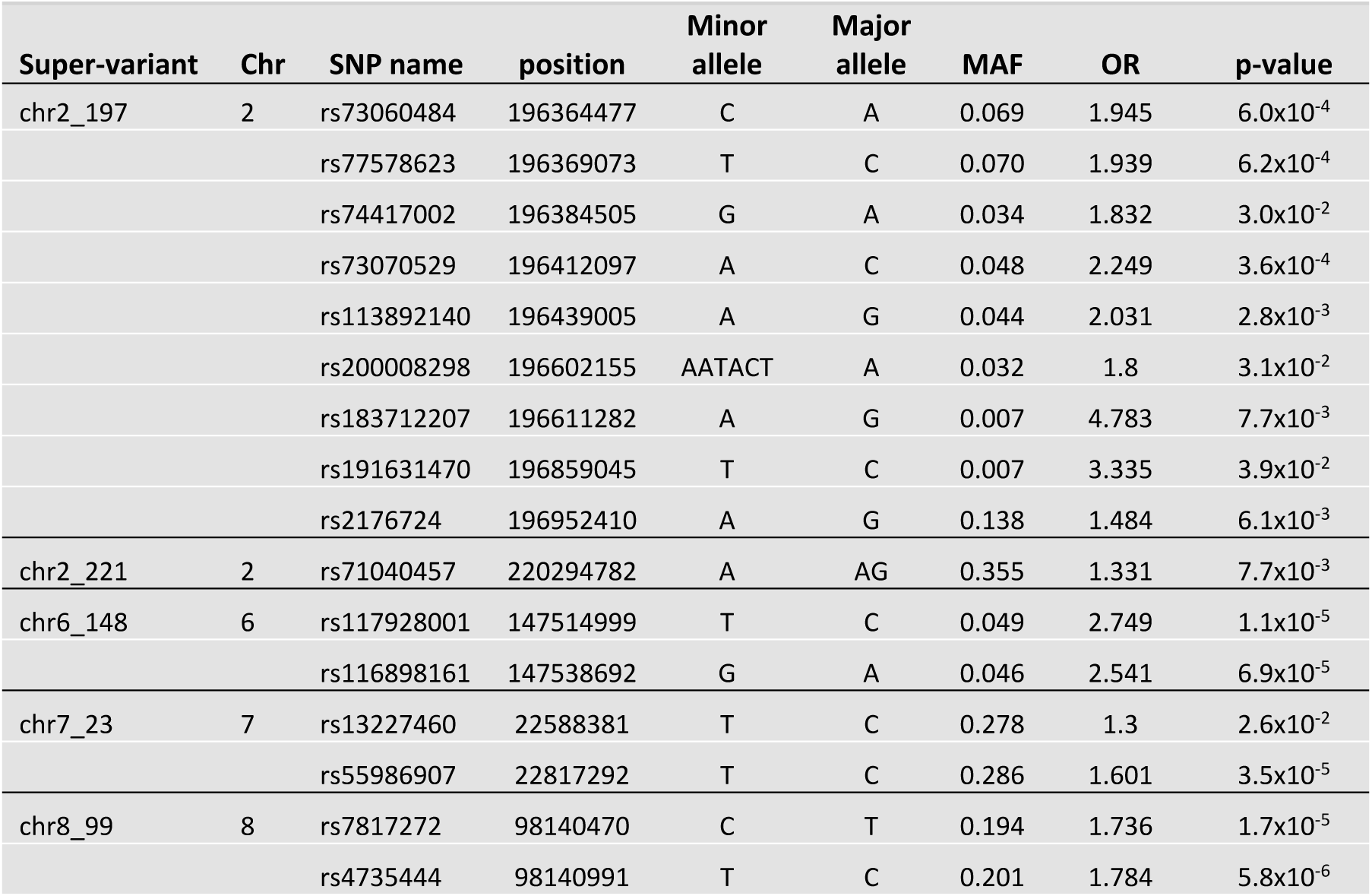
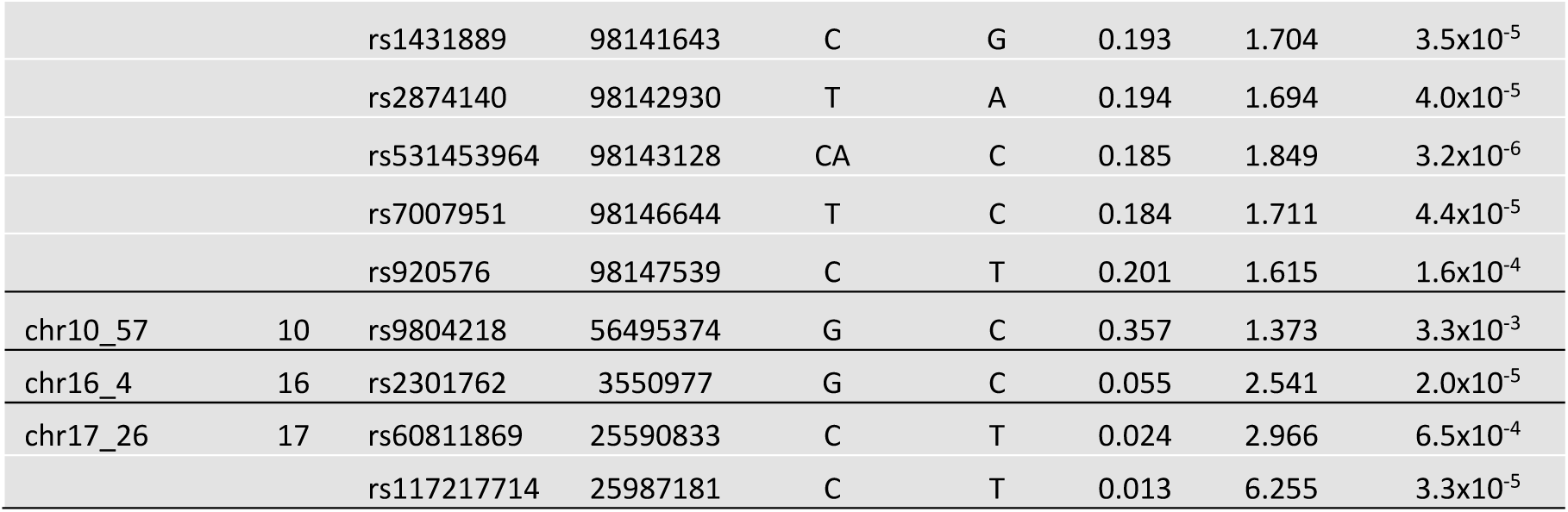
SNPs with high selection frequency and important gene mapping results in 8 super-variants.

**Figure 1:**
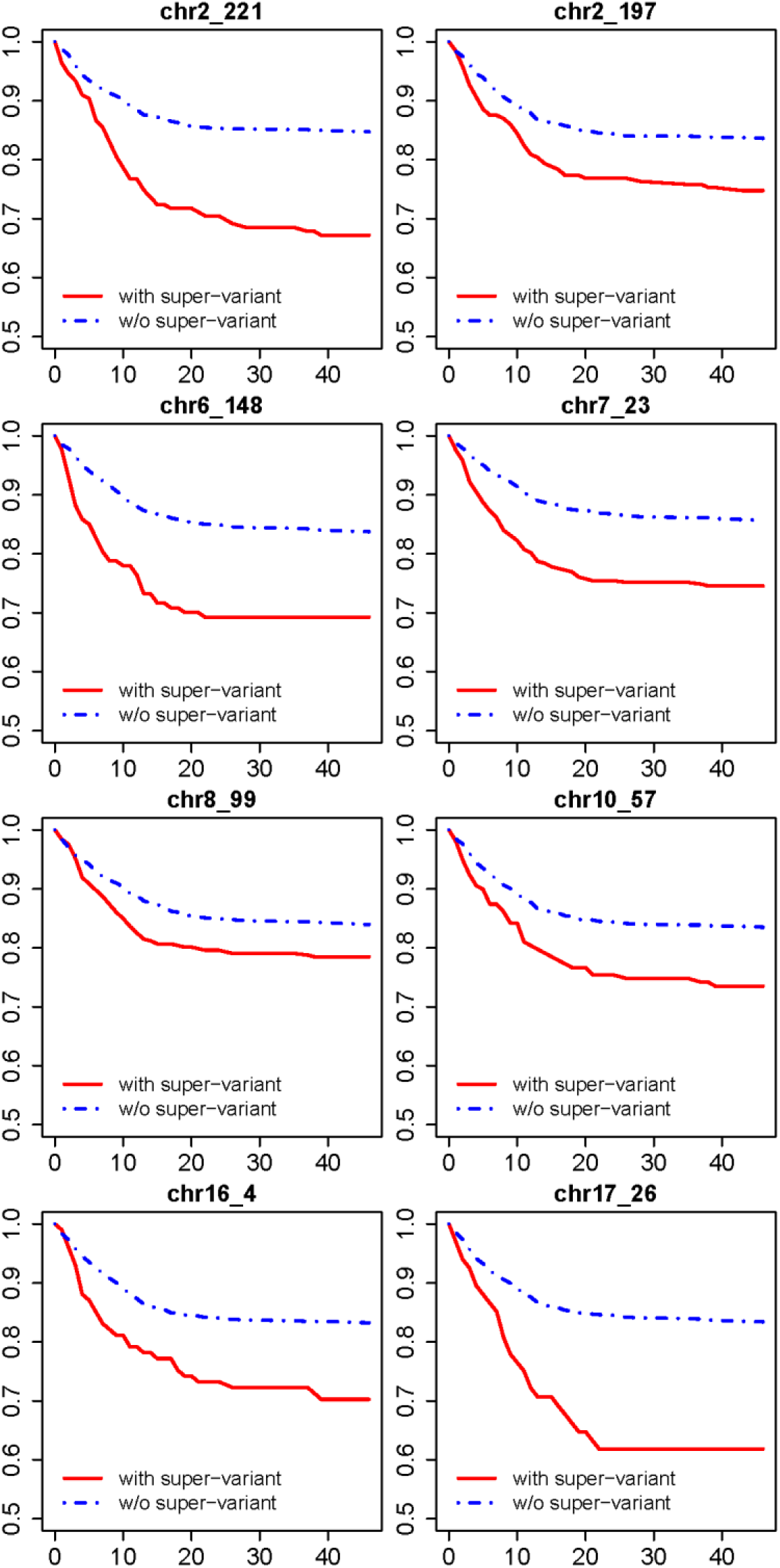
Survival curves of 8 identified super-variants in the complete dataset. The x-axis represents days since testing, and the y-axis represents the survival probability.

**Figure 2:**
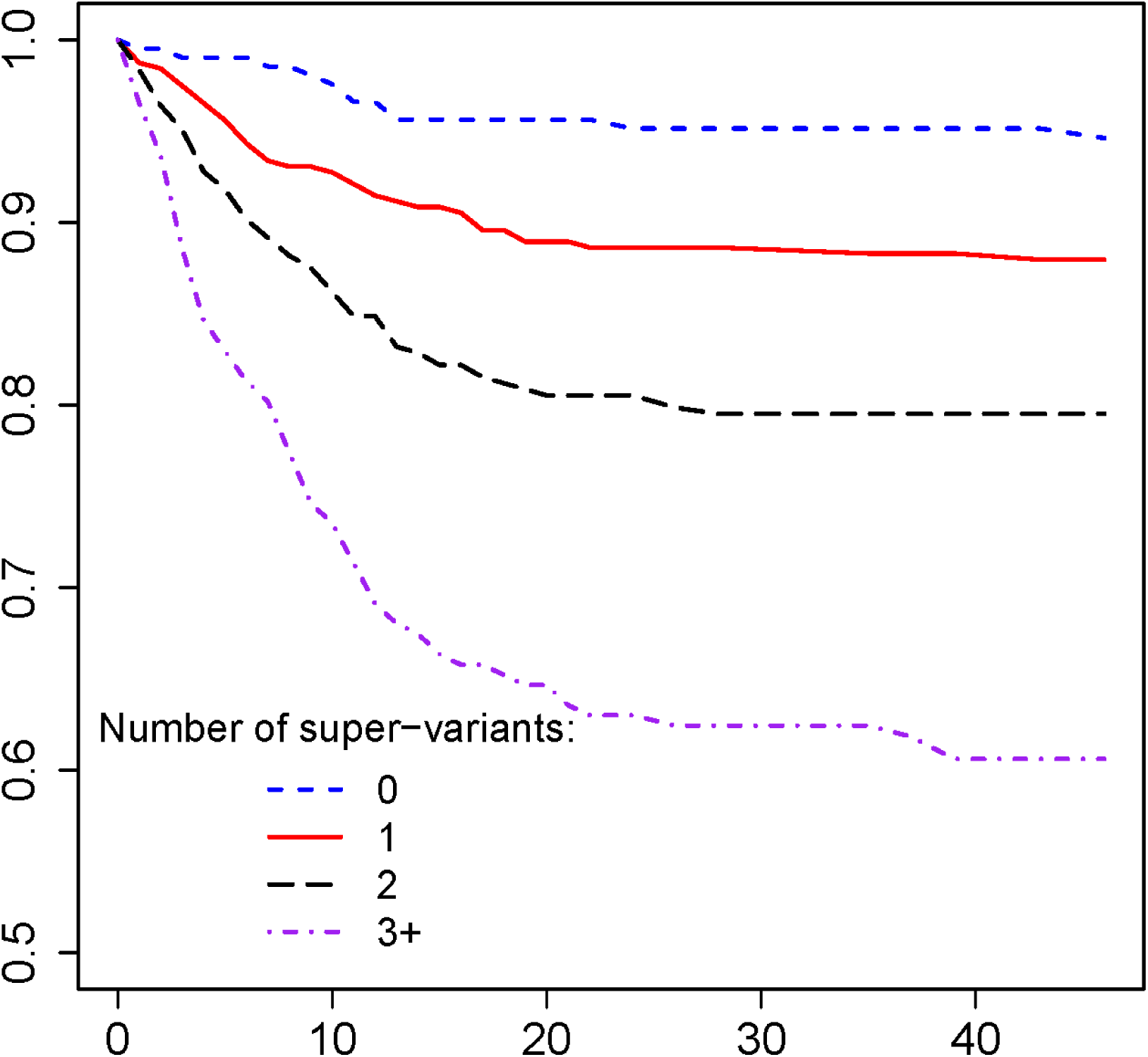
Survival curves stratified by the number of super-variants in the complete dataset. The x-axis represents days since testing, and the y-axis represents the survival probability.

In addition, we use a chi-square test for independence to investigate whether there are any gender differences among distribution of these 8 super-variants as well as differences among distribution of contributing SNPs. For super-variants, chr2_197 has p-value 0.0579 when all samples are considered. The frequency of presenting this super-variant among males and females is 18.09% and 22.93%, respectively. For contributing SNPs, rs4346407 on chromosome 2 has p-vale 0.050 when all samples are considered, and SNP 10:56525802_CT_C has p-value 0.0078 when only death cases are considered. The distributions of these two SNPs are given in Table 3.

**Table 3:**
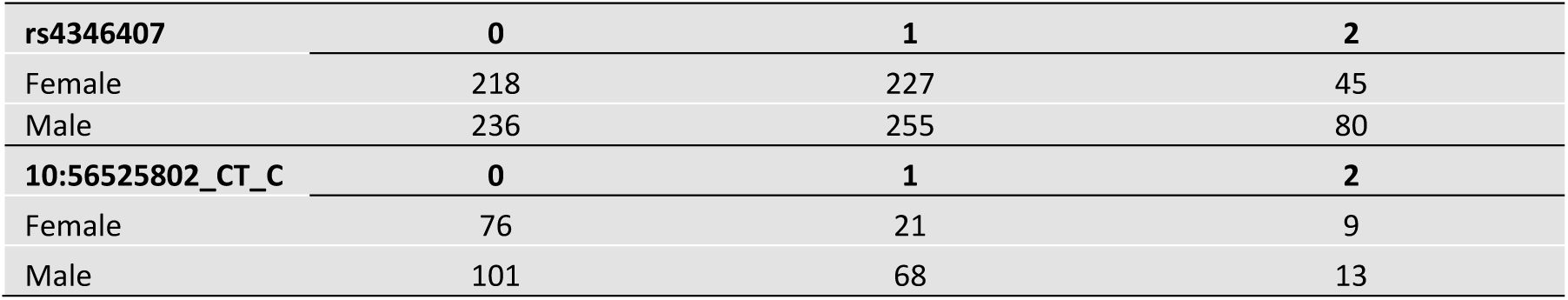
Allelic distribution of contributing SNPs.

## Discussion

As the COVID-19 pandemic creates a global crisis of overwhelming morbidity and mortality, it is urgent and imperative to provide insights into how host genetic factors link to clinical outcomes. With the timely release of UK Biobank COVID-19 dataset, we perform a GWAS study for detecting genetic risk factors for COVID-19 mortality. However, due to the limited sample size, the traditional single SNP GWAS has low power in signal detection which is evidenced by the Manhattan plot shown in Figure 3. This traditional association analysis is also conducted on the same samples with white British ancestry and controlled for gender and age. As demonstrated, the traditional single SNP analysis method is unable to detect any genome-wide significant association with commonly used threshold 5×10^−8^, which motivates us to consider the concept of super-variant for GWAS study.

**Figure 3:**
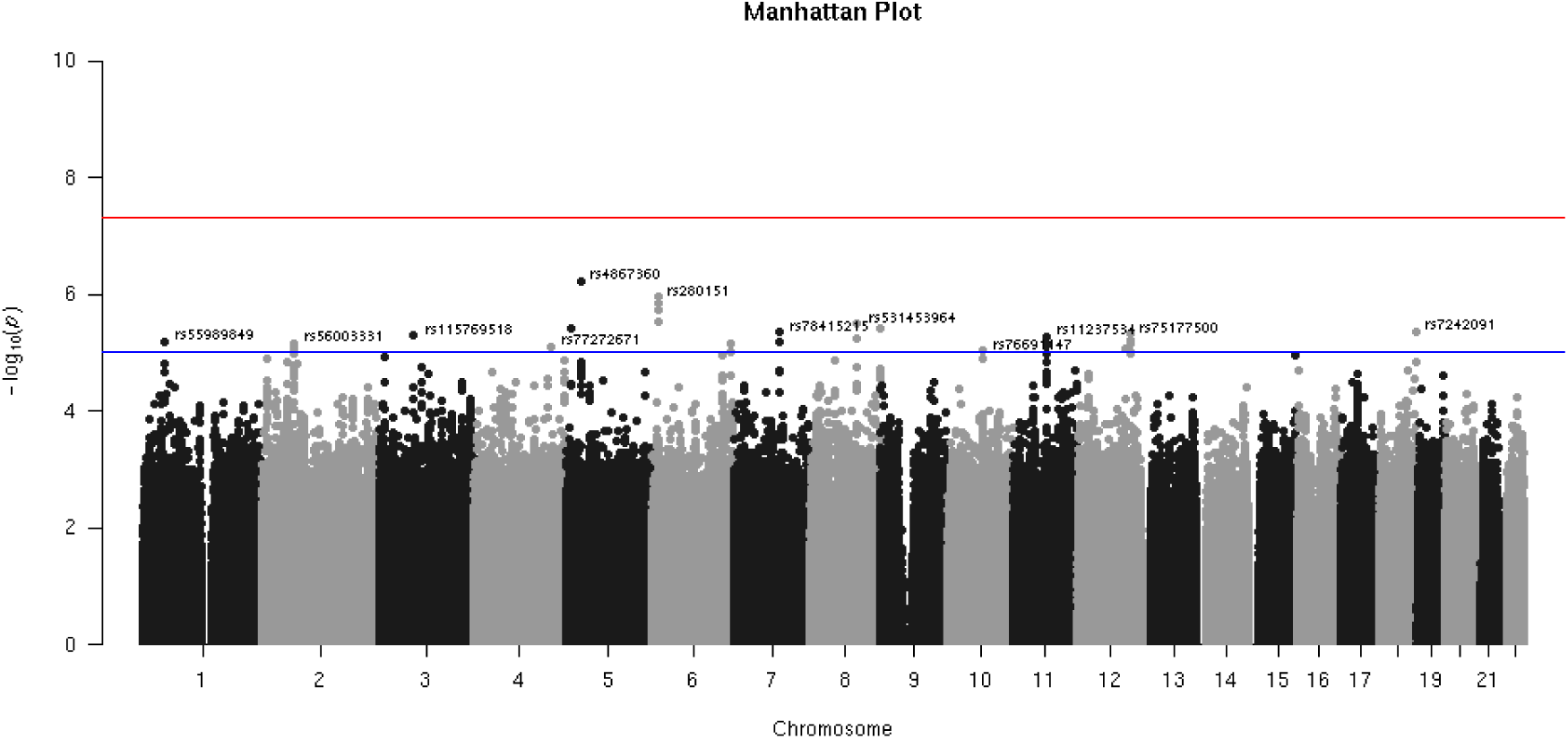
Manhattan plot of traditional single SNP association analysis based on samples with white British ancestry only and controlled for gender and age. The red horizontal line corresponds to the commonly adopted genome-wide significant level at 5×10^−8^, and the blue horizontal line gives to the suggestive significant level at 1×10^−5^. Top SNPs above the suggestive line in each chromosome are annotated.

Although the identified super-variants are similarly distributed in males and females, the results presented in Table 3 suggest that males tend to present more minor alleles for two contributing SNPs rs4346407 and 10:56525802_CT_C which potentially increase their risk of COVID-19 mortality. Such a phenomenon of higher risk for males has been reported in recent studies [17, 18, 26, 27].

The identified super-variants are mapped to annotated genes. The most interesting signal appears on chromosome 2 in the super-variant chr2_197. Within this super-variant, SNPs rs200008298, rs183712207, and rs191631470 are located in gene *DNAH7*. This gene encodes dynein axonemal heavy chain 7, which is a component of the inner dynein arm of ciliary axonemes. Gene Ontology (GO) annotations related to this gene include cilia movement and microtubule motor activity. A recently published paper showed that gene *DNAH7* is the most downregulated gene after infecting human bronchial epithelial cells with SARS-CoV2 [28]. The authors of that study speculated that the down-regulation of gene *DNAH7* causes the reduction of function of respiratory cilia. Our results suggest that COVID-19 patients with variations in gene *DNAH7* have higher risk for dying from COVID-19. We hypothesize that the disruption of *DNAH7* gene function may result in ciliary dysmotility and weakened mucociliary clearance capability, which leads to severe respiratory failure, a likely cause of COVID-19 death [29]. In addition, within the super-variant chr2_197, SNPs rs4578880 and rs113892140 are located in gene *SLC39A10*, which encodes a zinc transporter. This gene plays an important role in mediating immune cell homeostasis. It has been reported to facilitate antiapoptotic signaling during early B-cell development [30], modulate B-cell receptor signal strength [31], and control macrophage survival [32].

Signal at super-variant chr16_4 is also related to cilia. This super-variant consists of a single SNP rs2301762, which is located in gene *CLUAP1*. This gene encodes clusterin-associated protein 1. It is an evolutionarily conserved protein required for ciliogenesis [33], and its GO annotations include intraciliary transport involved in cilium assembly. Our findings evidence the importance of respiratory cilia functioning properly in COVID-19 patients, which may be an important site in host-pathogen interaction during SARS-CoV2 infection of airways [34] as well as a potential therapeutic target [35].

It is noteworthy that both super-variants chr2_197 and chr16_4 are related to cilia, which plays a crucial role in SARS-CoV-2 infection. Studies have reported that the angiotensin-converting enzyme II (ACE2) receptors on oral and nasal epithelium cells are the main portal for SARS-CoV-2 infection and transmission [36, 37]. Viral proliferation in the airway disrupts the structure and function of ciliated epithelium, causes ciliary dyskinesia and leads to lower respiratory tract infection [38]. Moreover, it has been reported that dysfunctions in olfactory cilia lead to loss of smell (anosmia), a COVID-19 associated symptom, and coronavirus hijacks the ciliated cells and causes deciliation in the human nasal epithelium [39].

Chr2_221 consists of 3 SNPs. SNP rs71040457 is located in the downstream of gene *DES* (distance = 3322 bp) and the upstream of gene *SPEG* (distance = 4917 bp). Gene *DES* encodes a muscle-specific class III intermediate filament. Its GO annotations include protein binding, structural constituent of cytoskeleton, and regulation of heart contraction. Gene *SPEG* encodes striated muscle enriched protein kinase, whose functions are related to protein kinase activity and muscle cell differentiation. Mutations in both gene *DES* and *SPEG* are reported to be associated with cardiomyopathy [40-42]. Several studies have reported cardiomyopathy in COVID-19 patients [43, 44], and acute myocardial damage caused by SARS-CoV-2 greatly increases the difficulty and complexity of patient treatment [45].

Chr7_23 is composed by five intergenic variant SNPs. Among them, SNP rs55986907 is an expression quantitative trait loci (eQTL) of gene *TOMM7* in multiple tissues, including whole blood, lung, adipose, thyroid, skin, nerve, and esophagus based on the Genotype-Tissue Expression (GTEx) database. The gene product of *TOMM7* is a subunit of the translocase of the outer mitochondrial membrane, and plays a role in regulating the assembly and stability of the translocase complex [46]. A study discussed that intra and extracellular mitochondrial function can be impacted by SARS-CoV-2, which may be related to the hyper-inflammatory state termed as the “cytokine storm” found in COVID-19 patients, with contributions to the progression and severity of the disease [47]. Super variant chr6_148 contains 101 SNPs. Eighty-nine of them are located in gene *STXBP5*and six of them are located in gene *STXBP5-AS1*. On the one hand, gene *STXBP5* encodes a syntaxin 1 binding protein. Its GO annotations include neurotransmitter release and regulation of synaptic vesicle exocytosis. Genome-wide association studies have found the association between *STXBP5* and Von Willebrand factor (VWF) plasma level in humans [48, 49], which is a predictor for the risk of myocardial infarction and thrombosis. A study showed that gene *STXBP5* inhibits endothelial exocytosis and promotes platelet secretion, and the variation within *STXBP5* is a genetic risk for venous thromboembolic disease [50]. COVID-19 leads to excessive inflammation, platelet activation, endothelial dysfunction, and stasis, which may predispose patients to venous and arterial thrombotic disease [51]. On the other hand, studies have revealed that *STXBP5-AS1* encodes a long noncoding RNA, which inhibits cell proliferation, migration, and invasion via preventing the phosphatidylinositol 3 kinase/protein kinase B (PI3K/AKT) signaling pathway against *STXBP5* expression in non-small-cell lung carcinoma and gastric cancer cells [52, 53]. Our results suggest that the variations within *STXBP5*/*STXBP5-AS1* and the interaction between them may result in increased risk of death among COVID-19 patients through the mechanism related to endothelial exocytosis.

Chr17_26 is composed by three intergenic variant SNPs. Among them, SNP rs60811869 is an eQTL of gene *WSB1* in Artery-Tibial tissue based on the GTEx database. Gene *WSB1* encodes a member of the WD-protein subfamily, which is highly expressed in spleen and lung [54]. Its related pathways include innate immune system and Class I MHC mediated antigen processing and presentation. This gene has been reported to function as a Lnterleukin-21(IL-21) receptor binding molecule, which enhances the maturation of IL-21 receptor [55]. Variations in this gene may result in disrupted functions of immune system and lead to higher death rate among COVID-19 patients.

Super-variant chr10_57 contains 11 SNPs and all of them are located in gene *PCDH15*. This gene is a member of the cadherin superfamily, which encodes a Calcium-dependent cell-adhesion protein. Gene *PCDH15* is essential for maintenance of normal retinal and cochlear function.

Super-variant chr8_99 is composed by 7 SNPs. All the SNPs are located in gene *CPQ*, which encodes carboxypeptidase Q. GO annotations of this gene include protein homodimerization activity and carboxypeptidase activity.

Although the roles of genes *PCDH15* and *CPQ* in viral infection remain largely unclear, our results warrant future investigation to learn the relationship between genetic variations and the severe COVID-19 outcomes.

Our study is restricted by the limited sample size. We anticipate a continuous accumulation of data in the following months and plan to iterate our analysis whenever more data become available. Furthermore, we currently focus on the population with white British ancestry of UK Biobank in the analysis, validating the identified risk factors in independent populations from other resources or ethnic groups worth further investigation.

## Conclusions

We identify 8 potential genetic risk loci for the mortality of COVID-19. These findings may provide timely evidence and clues for better understanding the molecular pathogenesis of COVID-19 and genetic basis of heterogeneous susceptibility, with potential impact on new therapeutic options.

## Data Availability

The data used in the study are available with the permission of the UK Biobank. 

https://www.ukbiobank.ac.uk

## Declarations

### Ethics approval and consent to participate

Ethical approval and participant consent were collected by UK Biobank at the time participants enrolled. This paper is an analysis of anonymized data provided by UK Biobank. According to Yale IRB, analysis of anonymized data does not constitute Human Subjects Research.

### Consent for publication

Not applicable.

### Availability of data and material

The data used in the study are available with the permission of the UK Biobank (https://www.ukbiobank.ac.uk).

## Authors’ contributions

JH, CL, and HZ designed the study. JH, CL, SW, and TL performed the experiments and analyzed the data. All authors made critical input to the manuscript.

## Acknowledgements

Zhang’s research is supported in part by U.S. National Institutes of Health (R01HG010171 and R01MH116527). This research has been conducted using the UK Biobank Resource under Application Number 42009. We thank the Yale Center for Research Computing for guidance and use of the research computing infrastructure.

