## Supplemental Table 1 for "Genetic variants are identified to increase risk of COVID-19 related mortality from UK Biobank data"

### Additional file 1

Additional file 1 contains a supplementary table.

**Table S1| SNPs corresponding to 8 super-variants. Statistics are based on complete dataset.**

| Super-variant | Chr | SNP name | position | Minor allele | Major allele | MAF | OR | p-value |
|---|---|---|---|---|---|---|---|---|
| chr2_197 | 2 | rs36142300 | 196091314 | C | A | 0.209 | 1.443 | $3.5x10^{-3}$ |
| | | rs78698341 | 196096984 | A | G | 0.163 | 1.367 | $2.4x10^{-2}$ |
| | | rs182681727 | 196112842 | T | C | 0.007 | 6.003 | $2.7x10^{-3}$ |
| | | rs4346407 | 196285029 | C | A | 0.345 | 1.13 | $2.7x10^{-1}$ |
| | | rs6434778 | 196289012 | C | T | 0.112 | 1.7 | $7.2x10^{-4}$ |
| | | rs7584942 | 196294552 | A | T | 0.176 | 1.397 | $1.3x10^{-2}$ |
| | | rs11677741 | 196295675 | A | G | 0.176 | 1.397 | $1.3x10^{-2}$ |
| | | rs13402652 | 196299017 | A | T | 0.176 | 1.388 | $1.5x10^{-2}$ |
| | | rs12614118 | 196304657 | T | C | 0.177 | 1.395 | $1.4x10^{-2}$ |
| | | rs10931697 | 196305450 | A | G | 0.110 | 1.728 | $5.2x10^{-4}$ |
| | | rs11674207 | 196305753 | G | A | 0.181 | 1.375 | $1.6x10^{-2}$ |
| | | rs10931698 | 196309354 | C | T | 0.177 | 1.392 | $1.4x10^{-2}$ |
| | | rs1542865 | 196309536 | C | A | 0.177 | 1.392 | $1.4x10^{-2}$ |
| | | rs16841170 | 196315854 | T | C | 0.068 | 1.925 | $7.8x10^{-4}$ |
| | | rs73058388 | 196316312 | T | C | 0.068 | 1.925 | $7.8x10^{-4}$ |
| | | rs73058392 | 196320779 | C | T | 0.068 | 1.974 | $4.7x10^{-4}$ |
| | | rs73058393 | 196322414 | A | G | 0.068 | 1.974 | $4.7x10^{-4}$ |
| | | 2:196329036_CT_C | 196329036 | C | CT | 0.068 | 1.963 | $5.2x10^{-4}$ |
| | | rs1500604 | 196330222 | G | A | 0.068 | 1.965 | $5.1x10^{-4}$ |
| | | rs111490104 | 196353285 | G | A | 0.069 | 1.952 | $5.6x10^{-4}$ |
| | | rs61554989 | 196356096 | G | C | 0.069 | 1.945 | $6.0x10^{-4}$ |
| | | rs6705003 | 196364147 | T | C | 0.069 | 1.945 | $6.0x10^{-4}$ |
| | | rs73060484 | 196364477 | C | A | 0.069 | 1.945 | $6.0x10^{-4}$ |
| | | rs56665249 | 196368539 | CA | C | 0.069 | 1.947 | $5.8x10^{-4}$ |
| | | rs74446597 | 196368733 | A | G | 0.069 | 1.898 | $9.2x10^{-4}$ |
| | | rs115471676 | 196369018 | C | G | 0.032 | 1.893 | $2.5x10^{-2}$ |
| | | rs77578623 | 196369073 | T | C | 0.070 | 1.939 | $6.2x10^{-4}$ |
| | | rs11885086 | 196372106 | A | G | 0.037 | 1.791 | $2.8x10^{-2}$ |
| | | rs78856931 | 196375644 | A | G | 0.032 | 1.964 | $1.6x10^{-2}$ |
| | | rs142172903 | 196379223 | G | A | 0.032 | 1.964 | $1.6x10^{-2}$ |
| | | rs74417002 | 196384505 | G | A | 0.034 | 1.832 | $3.0x10^{-2}$ |
| | | rs76910237 | 196386746 | C | T | 0.032 | 1.964 | $1.6x10^{-2}$ |
| | | rs115352051 | 196398982 | T | C | 0.032 | 1.964 | $1.6x10^{-2}$ |
| | | rs75314384 | 196400086 | G | C | 0.032 | 1.964 | $1.6x10^{-2}$ |
| | | rs73070529 | 196412097 | A | C | 0.048 | 2.249 | $3.6x10^{-4}$ |

| | | | | | | | | |
|---|---|---|---|---|---|---|---|---|
| | | rs144301033 | 196414150 | G | A | 0.032 | 1.964 | $1.6x10^{-2}$ |
| | | rs57201664 | 196418185 | C | G | 0.048 | 2.24 | $3.9x10^{-4}$ |
| | | rs112443640 | 196421676 | T | C | 0.048 | 2.24 | $3.9x10^{-4}$ |
| | | rs11896399 | 196425065 | T | C | 0.032 | 1.964 | $1.6x10^{-2}$ |
| | | rs11901900 | 196425141 | C | T | 0.032 | 1.964 | $1.6x10^{-2}$ |
| | | rs113892140 | 196439005 | A | G | 0.044 | 2.031 | $2.8x10^{-3}$ |
| | | rs4578880 | 196481450 | T | C | 0.162 | 1.196 | $1.9x10^{-1}$ |
| | | rs200008298 | 196602155 | AATACT | A | 0.032 | 1.8 | $3.1x10^{-2}$ |
| | | rs183712207 | 196611282 | A | G | 0.007 | 4.783 | $7.7x10^{-3}$ |
| | | rs191631470 | 196859045 | T | C | 0.007 | 3.335 | $3.9x10^{-2}$ |
| | | rs2176724 | 196952410 | A | G | 0.138 | 1.484 | $6.1x10^{-3}$ |
| | | 2:196973376_TGTATTCAATGA_T | 196973376 | T | TGTATTCAATGA | 0.007 | 6.029 | $2.3x10^{-3}$ |
| | | rs147853699 | 196995030 | C | T | 0.011 | 3.432 | $9.3x10^{-3}$ |
| chr2_221 | 2 | rs71040457 | 220294782 | A | AG | 0.355 | 1.331 | $7.7x10^{-3}$ |
| | | rs6711514 | 220828453 | T | C | 0.240 | 1.144 | $2.7x10^{-1}$ |
| | | rs34011564 | 220831743 | G | A | 0.240 | 1.152 | $2.4x10^{-1}$ |
| chr6_148 | 6 | rs117928001 | 147514999 | T | C | 0.049 | 2.749 | $1.1x10^{-5}$ |
| | | rs139385073 | 147517772 | C | G | 0.046 | 2.541 | $6.9x10^{-5}$ |
| | | rs58638359 | 147518533 | T | C | 0.046 | 2.541 | $6.9x10^{-5}$ |
| | | rs117813413 | 147519826 | A | T | 0.046 | 2.541 | $6.9x10^{-5}$ |
| | | rs112697374 | 147520440 | G | A | 0.046 | 2.541 | $6.9x10^{-5}$ |
| | | rs142112285 | 147520457 | T | G | 0.046 | 2.541 | $6.9x10^{-5}$ |
| | | 6:147522645_GGAAACTGGCTTAAATTGGTGACTCTTT_G | 147522645 | G | GGAAACTGGCTTAAATTGGTGACTCTTT | 0.041 | 2.468 | $3.5x10^{-4}$ |
| | | rs73788817 | 147528571 | A | G | 0.046 | 2.546 | $6.6x10^{-5}$ |
| | | rs117406405 | 147531562 | G | A | 0.046 | 2.541 | $6.9x10^{-5}$ |
| | | rs117668778 | 147532499 | G | C | 0.046 | 2.541 | $6.9x10^{-5}$ |
| | | 6:147535998_TA_T | 147535998 | T | TA | 0.046 | 2.556 | $6.1x10^{-5}$ |
| | | rs138108867 | 147536977 | G | A | 0.046 | 2.541 | $6.9x10^{-5}$ |
| | | rs117155560 | 147537958 | C | T | 0.046 | 2.541 | $6.9x10^{-5}$ |
| | | rs117620146 | 147538342 | G | T | 0.046 | 2.541 | $6.9x10^{-5}$ |
| | | rs116898161 | 147538692 | G | A | 0.046 | 2.541 | $6.9x10^{-5}$ |
| | | rs73788833 | 147541537 | G | A | 0.046 | 2.546 | $6.6x10^{-5}$ |
| | | rs118022104 | 147545290 | A | G | 0.046 | 2.546 | $6.6x10^{-5}$ |
| | | rs117739744 | 147546408 | G | A | 0.046 | 2.493 | $1.0x10^{-4}$ |
| | | rs148906599 | 147548941 | G | T | 0.045 | 2.433 | $1.7x10^{-4}$ |

| | | | | | | | |
|---|---|---|---|---|---|---|---|
| | rs79659451 | 147549365 | A | G | 0.045 | 2.438 | $1.7x10^{-4}$ |
| | rs117793403 | 147550879 | T | A | 0.045 | 2.438 | $1.7x10^{-4}$ |
| | rs17076671 | 147551381 | C | T | 0.045 | 2.438 | $1.7x10^{-4}$ |
| | rs118150890 | 147551986 | T | C | 0.045 | 2.438 | $1.7x10^{-4}$ |
| | rs144888374 | 147557853 | G | A | 0.045 | 2.438 | $1.7x10^{-4}$ |
| | rs149045958 | 147557993 | A | C | 0.045 | 2.438 | $1.7x10^{-4}$ |
| | rs9485155 | 147558268 | G | A | 0.045 | 2.438 | $1.7x10^{-4}$ |
| | 6:147558529_GT_G | 147558529 | G | GT | 0.045 | 2.431 | $1.8x10^{-4}$ |
| | rs79237089 | 147558600 | C | T | 0.045 | 2.438 | $1.7x10^{-4}$ |
| | rs73788845 | 147561055 | G | A | 0.045 | 2.438 | $1.7x10^{-4}$ |
| | rs568341244 | 147562842 | GTTA | G | 0.045 | 2.438 | $1.7x10^{-4}$ |
| | rs201745177 | 147562843 | TTA | T | 0.044 | 2.383 | $2.6x10^{-4}$ |
| | rs118031434 | 147563475 | A | C | 0.045 | 2.438 | $1.7x10^{-4}$ |
| | rs117826399 | 147563866 | G | T | 0.045 | 2.438 | $1.7x10^{-4}$ |
| | rs9497729 | 147564973 | C | A | 0.045 | 2.438 | $1.7x10^{-4}$ |
| | rs76364924 | 147565235 | G | A | 0.045 | 2.438 | $1.7x10^{-4}$ |
| | rs117483088 | 147565868 | T | A | 0.045 | 2.438 | $1.7x10^{-4}$ |
| | rs116841115 | 147567826 | G | C | 0.045 | 2.438 | $1.7x10^{-4}$ |
| | rs6921496 | 147568513 | C | T | 0.045 | 2.438 | $1.7x10^{-4}$ |
| | rs140618972 | 147569205 | T | C | 0.045 | 2.438 | $1.7x10^{-4}$ |
| | rs80096466 | 147570254 | C | T | 0.045 | 2.438 | $1.7x10^{-4}$ |
| | rs118119191 | 147574560 | T | C | 0.045 | 2.438 | $1.7x10^{-4}$ |
| | rs138150136 | 147575349 | A | G | 0.045 | 2.438 | $1.7x10^{-4}$ |
| | rs10499247 | 147577646 | A | C | 0.045 | 2.438 | $1.7x10^{-4}$ |
| | rs79378281 | 147579504 | T | C | 0.045 | 2.438 | $1.7x10^{-4}$ |
| | rs141169020 | 147579860 | T | C | 0.045 | 2.438 | $1.7x10^{-4}$ |
| | rs140169249 | 147579864 | T | C | 0.045 | 2.438 | $1.7x10^{-4}$ |
| | rs118000456 | 147580873 | A | G | 0.045 | 2.438 | $1.7x10^{-4}$ |
| | rs7743335 | 147584626 | C | T | 0.042 | 2.695 | $5.2x10^{-5}$ |
| | 6:147592734_TG_T | 147592734 | T | TG | 0.044 | 2.382 | $2.7x10^{-4}$ |
| | rs77337125 | 147599572 | G | A | 0.045 | 2.438 | $1.7x10^{-4}$ |
| | rs117920436 | 147599903 | A | C | 0.045 | 2.438 | $1.7x10^{-4}$ |
| | rs79747617 | 147600143 | G | A | 0.045 | 2.438 | $1.7x10^{-4}$ |
| | rs117486328 | 147603091 | T | C | 0.044 | 2.382 | $2.7x10^{-4}$ |
| | rs117810535 | 147603700 | C | T | 0.044 | 2.382 | $2.7x10^{-4}$ |
| | rs9485159 | 147605198 | A | C | 0.044 | 2.382 | $2.7x10^{-4}$ |
| | rs75775640 | 147605320 | C | T | 0.044 | 2.382 | $2.7x10^{-4}$ |
| | rs77912184 | 147606160 | G | A | 0.044 | 2.382 | $2.7x10^{-4}$ |
| | rs77553581 | 147609026 | C | A | 0.044 | 2.411 | $2.3x10^{-4}$ |

| | | | | | | | |
|---|---|---|---|---|---|---|---|
| | rs61120449 | 147609196 | T | C | 0.044 | 2.411 | $2.3x10^{-4}$ |
| | rs9497740 | 147610753 | A | G | 0.044 | 2.411 | $2.3x10^{-4}$ |
| | rs117405836 | 147612535 | G | A | 0.044 | 2.411 | $2.3x10^{-4}$ |
| | rs78796227 | 147612611 | C | T | 0.044 | 2.411 | $2.3x10^{-4}$ |
| | 6:147613781_GA_G | 147613781 | G | GA | 0.045 | 2.388 | $2.6x10^{-4}$ |
| | rs117170230 | 147614671 | A | C | 0.044 | 2.411 | $2.3x10^{-4}$ |
| | 6:147615018_TGAA_T | 147615018 | T | TGAA | 0.044 | 2.382 | $2.7x10^{-4}$ |
| | rs79107005 | 147615398 | G | T | 0.044 | 2.411 | $2.3x10^{-4}$ |
| | rs118152767 | 147619661 | T | A | 0.044 | 2.411 | $2.3x10^{-4}$ |
| | rs117009437 | 147619816 | T | C | 0.044 | 2.411 | $2.3x10^{-4}$ |
| | rs74966121 | 147621144 | G | A | 0.045 | 2.424 | $1.8x10^{-4}$ |
| | rs117942101 | 147621751 | A | C | 0.044 | 2.411 | $2.3x10^{-4}$ |
| | rs17076689 | 147622856 | C | G | 0.044 | 2.411 | $2.3x10^{-4}$ |
| | rs77234195 | 147623492 | T | C | 0.044 | 2.411 | $2.3x10^{-4}$ |
| | rs78617110 | 147632496 | T | G | 0.044 | 2.411 | $2.3x10^{-4}$ |
| | rs117453595 | 147633000 | A | C | 0.044 | 2.411 | $2.3x10^{-4}$ |
| | rs117583396 | 147637997 | A | G | 0.044 | 2.411 | $2.3x10^{-4}$ |
| | 6:147639198_TA_T | 147639198 | T | TA | 0.044 | 2.382 | $2.7x10^{-4}$ |
| | 6:147639804_CAG_C | 147639804 | C | CAG | 0.044 | 2.387 | $2.6x10^{-4}$ |
| | rs117535110 | 147643454 | T | A | 0.044 | 2.411 | $2.3x10^{-4}$ |
| | rs147458086 | 147644589 | A | G | 0.044 | 2.411 | $2.3x10^{-4}$ |
| | rs9497747 | 147647129 | C | T | 0.044 | 2.411 | $2.3x10^{-4}$ |
| | rs117235513 | 147647368 | T | C | 0.044 | 2.411 | $2.3x10^{-4}$ |
| | rs117463826 | 147648751 | A | G | 0.044 | 2.411 | $2.3x10^{-4}$ |
| | rs78684105 | 147649077 | A | G | 0.044 | 2.411 | $2.3x10^{-4}$ |
| | rs79211513 | 147649256 | G | A | 0.044 | 2.411 | $2.3x10^{-4}$ |
| | rs117704602 | 147649624 | A | C | 0.044 | 2.411 | $2.3x10^{-4}$ |
| | rs141785076 | 147650408 | C | T | 0.044 | 2.382 | $2.7x10^{-4}$ |
| | rs117136022 | 147651546 | C | G | 0.044 | 2.411 | $2.3x10^{-4}$ |
| | 6:147657277_GT_G | 147657277 | G | GT | 0.043 | 2.337 | $4.2x10^{-4}$ |
| | rs180680681 | 147659586 | T | C | 0.044 | 2.411 | $2.3x10^{-4}$ |
| | rs184939971 | 147659587 | C | T | 0.044 | 2.411 | $2.3x10^{-4}$ |
| | rs201971768 | 147660023 | GT | G | 0.041 | 2.284 | $8.6x10^{-4}$ |
| | rs117824912 | 147663694 | C | T | 0.044 | 2.411 | $2.3x10^{-4}$ |
| | rs117481329 | 147665615 | T | G | 0.044 | 2.411 | $2.3x10^{-4}$ |
| | rs79750405 | 147670272 | G | A | 0.045 | 2.408 | $2.2x10^{-4}$ |
| | rs117917771 | 147690430 | A | G | 0.044 | 2.411 | $2.2x10^{-4}$ |
| | rs78795351 | 147690643 | G | T | 0.044 | 2.411 | $2.2x10^{-4}$ |
| | rs41285881 | 147695051 | C | G | 0.044 | 2.411 | $2.2x10^{-4}$ |

| | | | | | | | | |
|---|---|---|---|---|---|---|---|---|
| | | rs58371914 | 147702098 | G | A | 0.044 | 2.411 | $2.2x10^{-4}$ |
| | | 6:147711840_TGA_T | 147711840 | T | TGA | 0.044 | 2.403 | $2.4x10^{-4}$ |
| | | rs117551998 | 147713497 | G | A | 0.044 | 2.331 | $4.2x10^{-4}$ |
| | | rs445693 | 147813931 | C | T | 0.018 | 2.226 | $2.3x10^{-2}$ |
| chr7_23 | 7 | rs1636305 | 22013049 | T | C | 0.495 | 1.213 | $6.5x10^{-2}$ |
| | | rs1637073 | 22013051 | C | T | 0.495 | 1.213 | $6.5x10^{-2}$ |
| | | rs7806543 | 22013935 | A | C | 0.483 | 1.2 | $8.5x10^{-2}$ |
| | | rs13227460 | 22588381 | T | C | 0.278 | 1.3 | $2.6x10^{-2}$ |
| | | rs55986907 | 22817292 | T | C | 0.286 | 1.601 | $3.5x10^{-5}$ |
| chr8_99 | 8 | rs7817272 | 98140470 | C | T | 0.194 | 1.736 | $1.7x10^{-5}$ |
| | | rs4735444 | 98140991 | T | C | 0.201 | 1.784 | $5.8x10^{-6}$ |
| | | rs1431889 | 98141643 | C | G | 0.193 | 1.704 | $3.5x10^{-5}$ |
| | | rs2874140 | 98142930 | T | A | 0.194 | 1.694 | $4.0x10^{-5}$ |
| | | rs531453964 | 98143128 | CA | C | 0.185 | 1.849 | $3.2x10^{-6}$ |
| | | rs7007951 | 98146644 | T | C | 0.184 | 1.711 | $4.4x10^{-5}$ |
| | | rs920576 | 98147539 | C | T | 0.201 | 1.615 | $1.6x10^{-4}$ |
| chr10_57 | 10 | rs9804313 | 56493670 | C | G | 0.352 | 1.366 | $3.5x10^{-3}$ |
| | | rs34974391 | 56494492 | T | G | 0.272 | 1.377 | $5.5x10^{-3}$ |
| | | rs9804218 | 56495374 | G | C | 0.357 | 1.373 | $3.3x10^{-3}$ |
| | | rs12777065 | 56495641 | T | G | 0.270 | 1.373 | $6.2x10^{-3}$ |
| | | rs4545447 | 56506954 | G | A | 0.272 | 1.33 | $1.2x10^{-2}$ |
| | | rs2384529 | 56507333 | G | A | 0.272 | 1.331 | $1.2x10^{-2}$ |
| | | rs201125676 | 56508905 | A | AAT | 0.271 | 1.304 | $2.0x10^{-2}$ |
| | | rs34638574 | 56511621 | C | T | 0.272 | 1.315 | $1.6x10^{-2}$ |
| | | 10:56525802_CT_C | 56525802 | C | CT | 0.198 | 1.343 | $2.0x10^{-2}$ |
| | | rs117422725 | 56886629 | T | G | 0.016 | 1.818 | $9.9x10^{-2}$ |
| | | rs10825470 | 56897213 | T | A | 0.234 | 1.288 | $4.2x10^{-2}$ |
| chr16_4 | 16 | rs2301762 | 3550977 | G | C | 0.055 | 2.541 | $2.0x10^{-5}$ |
| chr17_26 | 17 | rs60811869 | 25590833 | C | T | 0.024 | 2.966 | $6.5x10^{-4}$ |
| | | rs60849750 | 25672526 | A | C | 0.020 | 2.83 | $2.8x10^{-3}$ |
| | | rs117217714 | 25987181 | C | T | 0.013 | 6.255 | $3.3x10^{-5}$ |
